# Development of Restricted and Repetitive Behaviors from 2-19 and Internalizing Symptom Outcomes in a Longitudinal Study of Autism

**DOI:** 10.1101/2022.06.05.22275712

**Authors:** Nina Masjedi, Elaine B. Clarke, Catherine Lord

## Abstract

Using data from a well-characterized longitudinal cohort, this study examined trajectories of restricted and repetitive behaviors (RRBs), specifically repetitive sensorimotor (RSM), insistence on sameness (IS), and verbal RRBs, as measured by the Autism Diagnostic Interview, Revised (ADI-R) from ages 2 to 19. Additionally, this study investigated relationships between RSM and IS trajectories and parent- and self-report depression and anxiety symptoms in early adulthood. Group-based trajectory modeling and multilevel modeling were used to investigate change in RRB subtypes. RSM and IS behaviors generally decreased from 2 to 19, though some participants experienced increases in these RRB subtypes from ages 2 to 9. 65% of this sample had sufficient verbal ability prior to age 19 to calculate trajectories of verbal RRBs. Of this subset, 49% had few to no verbal RRBs throughout development; in contrast, the remaining 51% experienced increasing verbal RRBs from 2 to 9, followed by a plateau in verbal RRBs from 9-19. Higher ADOS Social Affect (SA) CSS scores in early childhood were linked with more RSM symptoms across development, but not related to the IS and Verbal trajectories. Non-verbal IQ from early childhood was not connected to change in any of the identified RRB trajectories. There were no associations between IS trajectories and internalizing symptom in early adulthood. However, preliminary data suggests that a Moderate-Decreasing pattern of RSM development may be linked to anxiety in early adulthood. These findings illustrate continuity and change in a core ASD symptom domain, RRBs, from early childhood to early adulthood.

## Introduction

Autism spectrum disorder (ASD) is a lifelong condition. The behavioral phenotype of ASD is not static; symptoms change across development. Symptoms of ASD are typically conceptualized under two core categories: social communication and restricted and repetitive behaviors (RRBs). Social communication impairments in ASD include deficits in eye contact, limited use of facial expressions and gestures, and difficulty maintaining relationships (American Psychiatric Association, 2014). RRBs include stereotyped motor movements, repetitive use of objects, and unusual sensory interests (APA, 2014). Importantly, RRBs vary considerably both across and within individuals. Understanding changes in ASD symptom presentation across the life course may facilitate the design and use of targeted treatment approaches for individuals with ASD at all stages of development.

Prior work in the current sample—a well-characterized longitudinal study of individuals with ASD and individuals with non-spectrum developmental delays referred to ASD clinics in early childhood—examined trajectories of social communication deficits from ages 2 to 19 and found significant improvements in social communication with increasing age (Bal et al., 2019). Social communication improved during this period in participants of all verbal abilities, though verbally fluent participants experienced greater improvements in social communication than minimally-verbal participants and participants with delayed speech (Bal et al., 2019). Another study using the current longitudinal sample of ASD examined trajectories of RRBs—specifically Repetitive Sensory Motor (RSM) and Insistence on Sameness (IS) behaviors—from ages 2 to 9 (Richler et al., 2010). Participants’ RSM and IS behaviors were analyzed separately, then compared by diagnostic category (i.e., autism spectrum disorder, pervasive developmental disorder, not otherwise specified [PDD-NOS], or non-autism developmental delays). Amongst participants with an ASD diagnosis, Richler et al. found three distinct trajectory groups for RSM scores, a mild group, a slightly decreasing group, and a consistently severe group, as well as three groups for IS scores, a mild group, an increasing group, and a moderate group. Whereas RSM scores had consistent severity or improved slightly from ages 2-9, IS scores remained consistent or increased during this period. In line with Richler et al.’s previous findings, the current study only examines RRB trajectories in participants in the longitudinal sample with a clinical diagnosis of ASD.

The prior studies of the current longitudinal sample used *a priori* analytic groupings based on participant language level (Bal et al., 2019) and diagnostic status (Richler et al., 2010) to identify trajectories of ASD symptoms. In contrast, this paper builds upon previous work in the same sample to examine continuity and change in ASD symptomatology across the lifespan by 1) analyzing RRB trajectories from ages 2-19 and 2) using a data-driven approach to identify participant trajectory groupings.

### Developmental Characterizations of RRBs

Several widely-available diagnostic instruments can be used to measure RRBs, notably the Autism Diagnostic Interview-Revised (ADI-R; Rutter et al., 2003) the Autism Diagnostic Observation Schedule, 2^nd^ Edition (ADOS-2;Lord et al., 2012) and the Repetitive Behavior Scale-Revised (RBS-R; Lam, 2004). In line with the prior longitudinal analysis of RRBs in this sample (Richler et al., 2010), the current study used clinician ratings from the ADI-R to assess change in RRBs across development. A separate cross-sectional study using the Simons Simplex Collection (SSC; n = 1,825), suggested the ADI-R can capture subcategories of RRBs in more detail than what can typically be observed in an ADOS-2 administration (Bishop et al., 2013). In contrast to the RBS-R, the ADI-R has the additional strength of being a clinical interview rather than a questionnaire, and is therefore less apt to be skewed by reporting bias (McDermott et al., 2020; Mirenda et al., 2010).

Previous factor analyses of the ADI-R have found two emergent factors of RRBs: Repetitive Sensorimotor (RSM) and Insistence on Sameness (IS) behaviors (Bishop et al., 2013, Richler et al., 2010). RSM includes behaviors such as hand and finger mannerisms and complex body mannerisms, repetitive use of objects, and unusual sensory interests. IS includes behaviors such as compulsions and rituals, difficulties with changes in routine, and resistance to trivial changes in the environment (Cuccaro et al., 2003; Richler et al., 2007, 2010). Separate factor analyses of both the ADI-R and the RBS-R have identified RSM and IS factors, suggesting these subcategories are not a product of the structure of the ADI-R (Cuccaro et al., 2003; Richler et al., 2010; Szatmari et al., 2006). Notably, other studies of the factor structure of ADI-R have identified additional RRB factors but of single items including stereotyped speech (Hiruma et al., 2021) and circumscribed interests (Lam et al., 2008).

Though few studies have examined RRBs longitudinally from early childhood to early adulthood, prior work in separate samples has examined changes in RRBs across childhood. Similar to Richler et al., (2010), these studies have found that many RRB behaviors increase from early to mid-childhood (Courchesne et al., 2021; Guthrie et al., 2013; Moore & Goodson, 2003). In contrast, cross-sectional studies of adults with ASD indicate RSM behaviors decrease with increasing age (Bishop et al., 2013; Esbensen et al., 2009; Evans et al., 2017; Uljarević et al., 2021). Separate Cross-sectional studies have found that IS behaviors increase with increasing age (Gotham et al., 2013). Considered as a whole, the literature on RRBs in childhood and early adulthood suggest RRB trajectories before and after late childhood may follow differing slopes, with RRBs increasing through mid-to late-childhood, and decreasing into adolescence and early adulthood. Additionally, these slopes may vary by subtypes of RRBs, though there is currently not sufficient evidence to establish this. The present study specifically examines change in the slopes of RRB trajectories, and tests whether these slopes significantly differ before and after mid-childhood.

### Verbal RRBs

Verbal ability and communicative use of language vary amongst people with autism and are a key predictor of developmental trajectories (Magiati et al., 2014; Pickles et al., 2020). It seems likely that language level could also play an important role in RRB trajectories. Some RRBs, such as verbal rituals and neologisms/idiosyncratic language, cannot be present in nonverbal autistic individuals. Other RRBs, such as circumscribed interests and unusual preoccupations, may be present in autistic individuals of varying language abilities but may be more obvious in verbally fluent individuals who can speak about their interests and preoccupations at length. In this study, verbal RRBs are defined as behaviors that are at least partially contingent on verbal ability, such as verbal rituals and circumscribed interests (see Table 1). For example, verbal rituals require some level of language fluency as defined in the ADI-R (Rutter et al, 2003) and circumscribed interests may be more apparent if a child is able to verbalize these interests.

**Table 1.** ADI-R Raw Items Include in RSM, IS, and Verbal Trajectory Sums.

| RRB (Total) | Trajectory |  |  | ADI-R Item |
| --- | --- | --- | --- | --- |
|  | RSM | IS | RRB (Verbal) |  |
| ? | ? |  |  | *Repetitive Use of Objects* |
| ? | ? |  |  | *Unusual Sensory Interests* |
| ? | ? |  |  | *Hand and Finger Mannerisms* |
| ? | ? |  |  | *Other Complex Mannerisms* |
| ? |  | ? |  | *Resistance to Trivial Changes in Environments* |
| ? |  | ? |  | *Difficulties with Changes in Routine* |
| ? |  | ? |  | *Compulsions and Rituals* |
| ? |  |  | ? | *Verbal Rituals* |
| ? |  |  | ? | *Unusual Preoccupations* |
| ? |  |  | ? | *Circumscribed Interests* |
| ? | □ |  | ? | *Stereotyped Utterances and Delayed Echolalia* |
| ? |  |  | ? | *Neologisms/Idiosyncratic Language* |

There is some ambiguity as to how to classify restricted and repetitive behaviors. For example, symptoms such as stereotyped utterances and delayed echolalia and neologisms/idiosyncratic language are not included in the RRB algorithm of the ADI-R (Rutter et al., 2003). In the original ADI interview, autism was diagnosed via symptoms in three components: social, RRBs, and communication, with this later domain encompassing verbal symptoms (Le Couteur et al., 1989). However, the current DSM-5 conceptualization of ASD stratifies symptoms into two domains—social communication deficits and RRBs—and communication symptoms are now included in the RRB diagnostic domain (APA, 2014). Trajectories of verbal RRBs have not been previously studied in detail (Volden & Lord, 1991). To quantify the influence of verbal ability on RRB trajectories, this study examines trajectories of verbal RRBs separately from trajectories of RSM and IS behaviors. For RSM and Verbal RRBs specifically, we also compare DSM-5 RRB criteria to the ADI-R algorithm criteria.

### RRBs and Internalizing Symptoms

There is some evidence that internalizing symptoms—such as anxiety and depression—in people with autism may be related to RRBs. People with autism are more likely to have high levels of internalizing symptoms compared to typical peers (Hollocks et al., 2018; Hu et al., 2019; Lai et al., 2019; Lugo-Marín et al., 2019; van Steensel & Heeman, 2017). RRBs may contribute to internalizing symptoms in people with autism. For example, the presence of RRBs may contribute to feelings of loneliness, as some RRBs, such as circumscribed interests and complex body mannerisms, may cause people with autism to be misunderstood or shunned by typical peers. One study found children with more severe restricted and repetitive behaviors were less connected with peers (Jones et al., 2017). The link between RRBs and decreased social connection could contribute to feelings of loneliness, and by extension, the development of depressive symptoms (Hedley et al., 2018). Further, IS is theoretically similar to intolerance of uncertainty, which is associated with anxiety and depression in both typical and neurodiverse populations (Boulter et al., 2014; Carleton et al., 2012).

Previous research has found associations between IS and anxiety, as well as associations between IS and depressive symptomology (Baribeau et al., 2021a; Gotham et al., 2013, 2014; Lidstone et al., 2014; Rodgers et al., 2012; Stratis & Lecavalier, 2013). In contrast, there have been few examinations of the association between RSM behaviors and internalizing symptoms. The relationship between RSM and depression has not been studied, and there is mixed evidence regarding RSM behaviors’ association with anxiety (Lidstone et al., 2014; Rodgers et al., 2012; Wurzman et al., 2015). However, several studies have found total RRBs are associated with both depression *and* anxiety (Baribeau et al., 2020; Lam, 2004; Stratis & Lecavalier, 2013). The current study sought to replicate prior findings of positive associations between IS behaviors and internalizing symptoms and to examine associations between RSM behaviors and internalizing symptoms.

### Study Aims

Our first aim was to examine trajectories of raw ADI-R RSM scores and raw ADI-R IS scores from ages 2 to 19. In line with prior findings in this sample, we expect ADI-R raw RSM scores to decrease and raw IS scores to increase with increasing age (Bishop et al., 2013; Esbensen et al., 2009; Evans et al., 2017; Guthrie et al., 2013; Moore & Goodson, 2003; Richler et al., 2010).

Our second aim was to examine the influence of verbal ability on RRBs from early childhood to early adulthood by identifying trajectories of ADI-R raw verbal RRB scores from ages 2 to 19.

Finally, our third aim was to characterize associations between ADI-R raw IS score and raw RSM score trajectory groups and internalizing symptoms in adulthood. In line with the existing literature, we expected RSM and IS behaviors would be associated with increased internalizing symptoms in early adulthood. (Baribeau et al., 2021; Gotham et al., 2013, 2014; Lidstone et al., Stratis & Lecavalier, 2013).

## Method

### Participants

193 consecutive referrals to community-based developmental clinics in North Carolina, Chicago, and Michigan were recruited to a longitudinal study of autism. All participants had been given a diagnosis of ASD, and completed the ADI-R at least once *(m* = 3.9 visits; SD = 1.2). Of the 193 participants, 154 individuals had completed at BDI, AMAS, or CBCL Parent Report Data collection at least once from ages 9 through 26. Attrition occurred as the result of refusal, change of place of residence, and inaccessible status. Participants lost due to attrition were more likely to have a mother with less than a college degree, *X**^2^***(1, 193) = 5. 73, *p* = 0.016. The descriptive characteristics of the sample are shown in Table S1.

### Procedure

Face-to-face visits occurred at approximately ages 2, 3, 5, 9, and 19. Research-reliable research assistants, clinical psychologists, or trainees administered the ADI-R and other diagnostic measures. Caregivers and, when capable, participants also completed demographic questionnaires and measures of internalizing symptomatology. Parents and participants over the age of 18 who were their own legal guardians signed a consent form as required by the applicable institutional review board(s) prior to each visit.

### Measures

#### Autism Symptomology

The ADI-R (Rutter et al., 2003) was administered at ages 2, 3, 5, 9, and 19. The ADI-R was used to calculate verbal RRB trajectories, RSM trajectories, and IS trajectories using the following time points: ages 2, 3, 5, 9, 19. The ADI-R items used to calculate each trajectory are shown in Table 1. To capture nuanced changes in RRB subtypes, the raw score (0 through 3, with 3 meaning most severe) for each item was used in a total sum for the domain. If any less than all items in a domain were missing, then those items were filled with a zero. If all items in a domain were missing, a score for that domain was not calculated. The domains reflect higher severity of symptoms as the sum increases. For the verbal RRB trajectories, only participants with a score of 0 for the ADI-R item “Overall Level of Language,” at age 19 or 9 were included in the trajectory analysis.

#### IQ

At each face-to-face assessment, verbal and non-verbal IQ were calculated using one of the following assessments (arranged here from most to least difficult): the Wechsler Abbreviated Scale of Intelligence (Wechsler, 1999), the Differential Ability Scales (Elliott, 2007), or the Mullen Scales of Early Learning (Mullen, 1995). Further details on the selection of appropriate IQ measures in this sample are described in prior work (Anderson et al., 2014; Lord et al., 2006).

#### Internalizing Symptoms

The Beck Depression Inventory, Second Edition (BDI-II; Beck et al., 1996) and the Adult Manifest Anxiety Scale (AMAS; Lowe & Reynolds, 2004) were given to parents and participants who could read at a third-grade level at face-to-face visits at two time points in young adulthood, once at approximately age 19 (*m* = 18.59, SD = 1.46) and again seven years later, at approximately age 26 (*m* =25.53, SD = 1.41). Parent- and self-report AMAS-A Total Anxiety scores (Reynolds et al., 2003) and BDI overall total scores (Beck et al., 1996) were used in the current analyses.

Prior analyses of this sample (McCauley et al., 2020) established separate trajectories of anxiety and depression symptoms from ages 9-26 based on parent- and self-report Anxious/Depressed T scores from the Child Behavior Checklist (CBCL; Achenbach & Rescorla, 2001) and Adult Behavior Checklist (ABCL; Achenbach & Rescorla, 2003). McCauley et al. found that participants’ anxiety symptoms were characterized by either a Stable-Low or a Stable-High trajectory over time, and depressive symptoms followed either Stable-Lor or High-Fluctuating trajectories. The likelihood of inclusion in these depression and anxiety trajectory groups was compared across the IS and RSM trajectory groups established in the current study.

### Data Analysis

To examine trajectories of current ADI-R RRB scores, traj plugin in Stata 16 was used to perform group-based trajectories. Group-based trajectory modeling estimates developmental trajectories via maximum likelihood estimation, and missing data are handled by estimating the model using all available information. The best fitting model (linear, quadratic, etc.) and number of trajectory groups were determined using Bayesian Information Criteria (BIC). Five time points were used as independent variables to analyze the latent grouping of individuals in their RRB trajectories. Unconditional 2, 3, and 4 class models were compared using Bayesian Information Criterion (BIC) and the smallest group membership percentage (Figure 1; Figure 2; Figure 3). After classes were determined, higher order effects were tested to establish whether cubic, quadratic, linear, or intercept modeling best explained variation over time. To aid in model selection, the average posterior probabilities were evaluated to determine adequate model fit (above 0.70; Nagin et al., 2018). To assess potential variation in the trajectories of specific types of RRBs, RSM score trajectories— which included all ADI-R C-section items assessing repetitive sensorimotor behaviors—IS score trajectories—which included all ADI-R C-section items assessing insistence on sameness behaviors—and verbal RRBs trajectories—which included all ADI-R C-section items section items at least partially contingent on verbal ability—were separately calculated. The ADI-R items included in each of the three symptom domain trajectories are summarized in Table 1.

**Figure 1.**
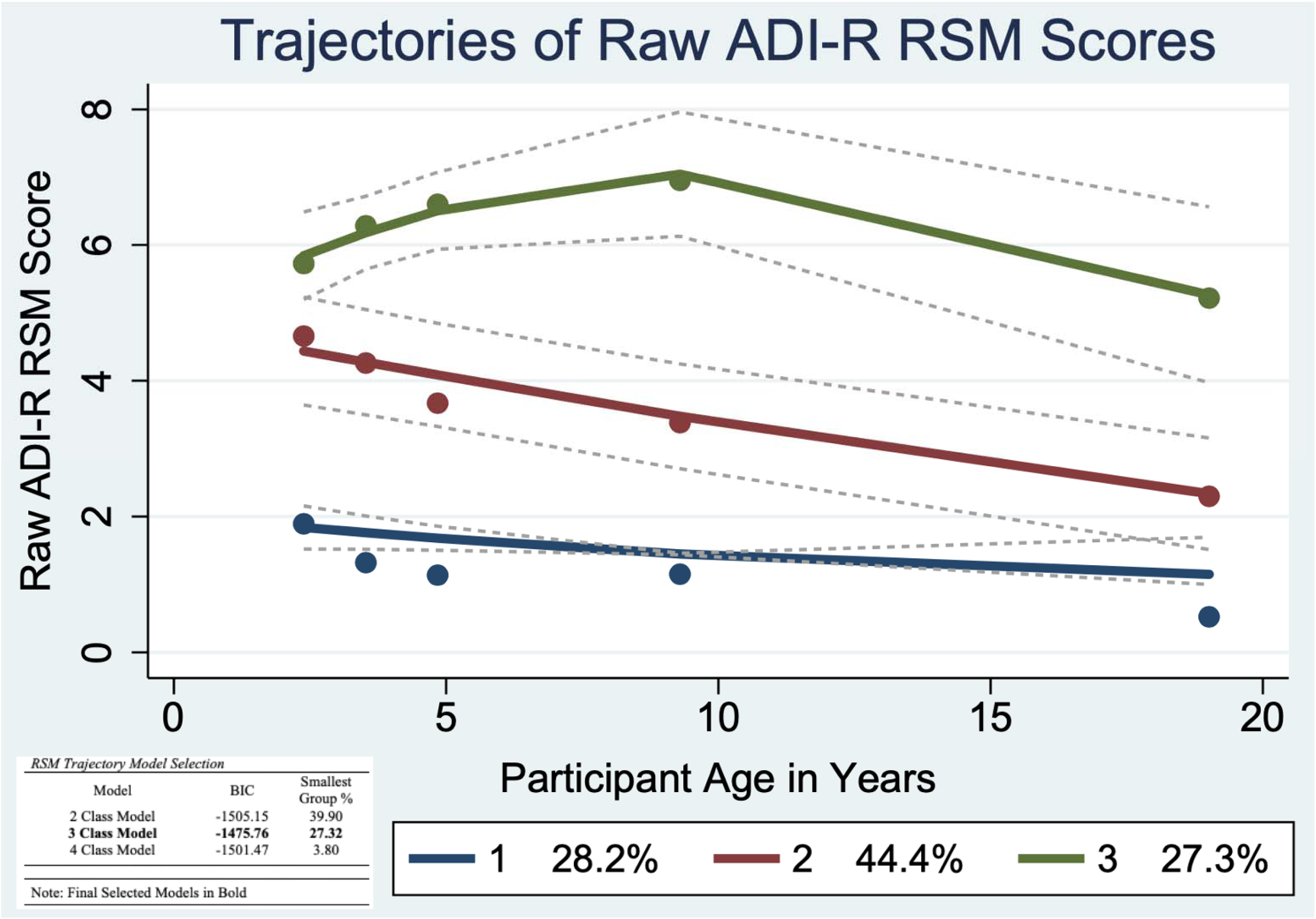
Trajectories of Raw ADI-R Restricted Sensorimotor (RSM) Scores

**Figure 2.**
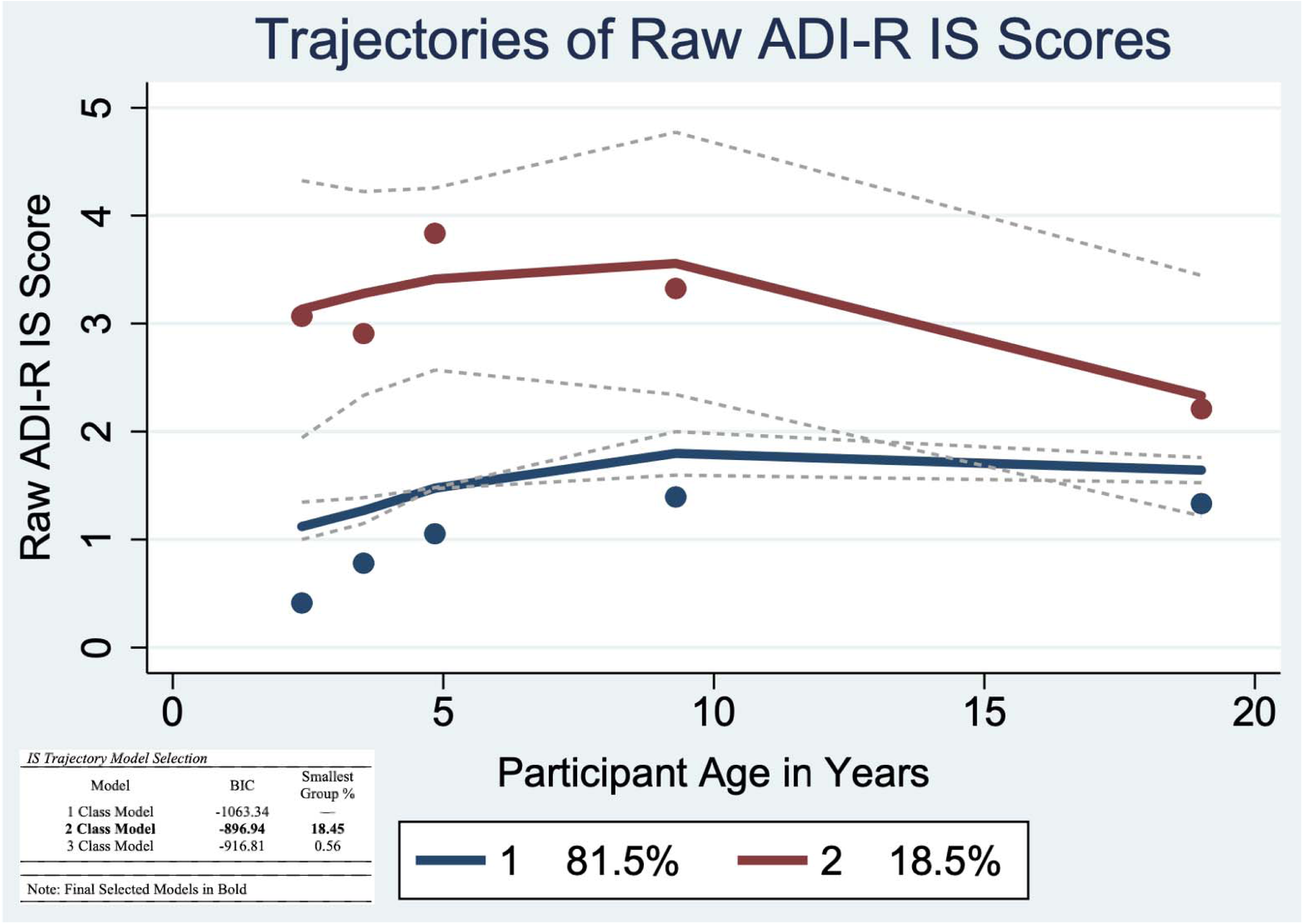
Trajectories of Raw ADI-R Insistence on Sameness (IS) Scores

**Figure 3.**
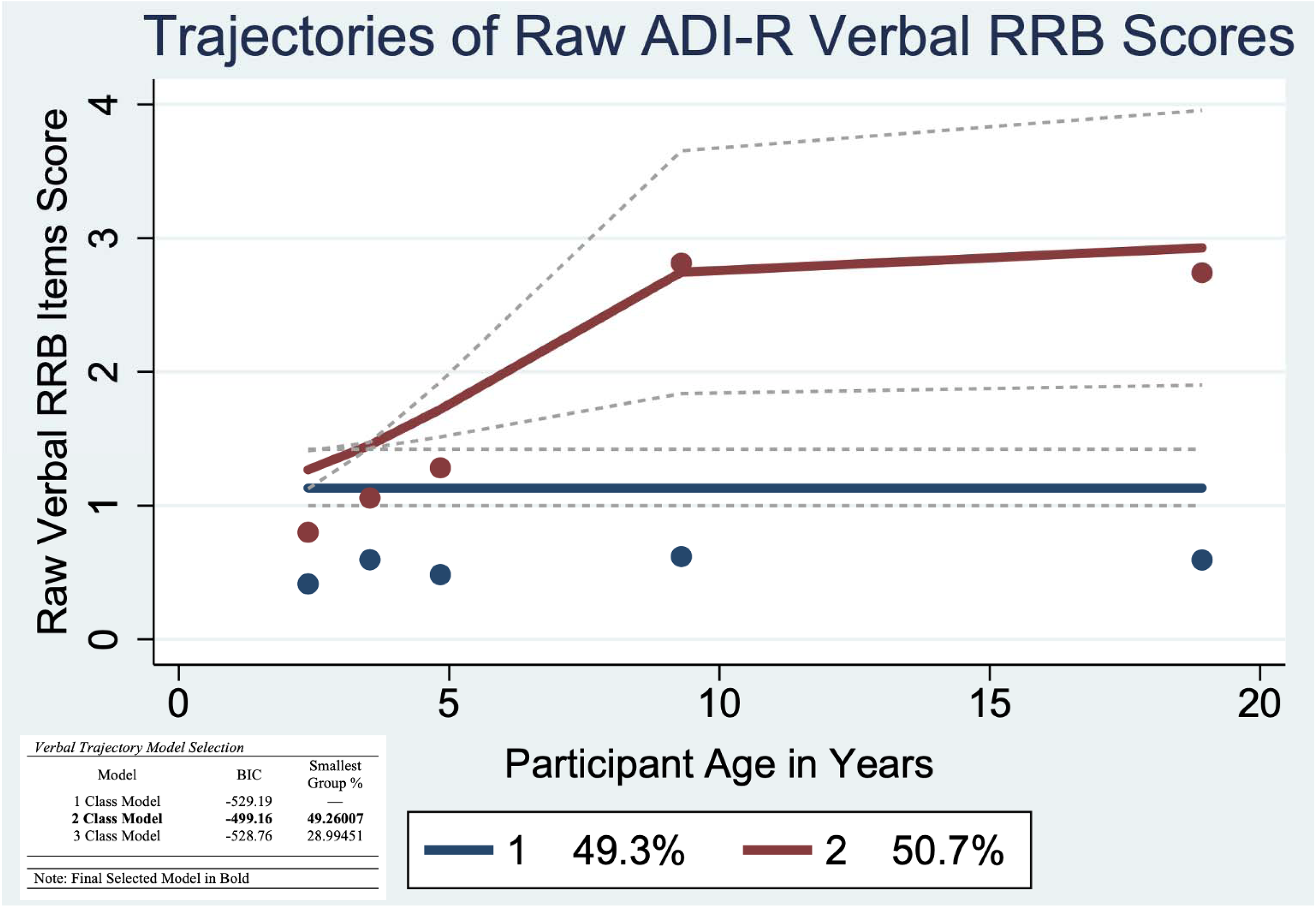
Trajectories of Raw ADI-R Verbal Restricted Repetitive Behavior (RRB) Scores

To account for the inclusion of communication symptoms in the RRB domain of DSM-5 ASD diagnostic criteria (APA, 2014), we also calculated totals for RSM trajectories that included the raw item scores for stereotyped utterances and delayed echolalia, as well as totals for verbal trajectories that included the raw item scores for stereotyped utterances, delayed echolalia and neologisms/idiosyncratic language. The inclusion of these items in the RSM and verbal trajectories did not demonstrably change the trajectory group membership percentages or patterns of growth (see Figure S3). Thus, we proceeded with the original domains based on the ADI-R algorithms and prior factor analyses, rather than DSM-5 diagnostic criteria.

One-way ANOVA and chi-square analyses were used to compare the composition of the RSM (Table 2), IS (Table 3), and Verbal (Table 4) trajectory classes for gender, race, recruitment site, caregiver education, childhood ADOS CSS scores and IQ, parent- and self-report BDI and AMAS scores, and inclusion in the previously established depression and anxiety trajectory groups (McCauley et al., 2020).

**Table 2.** Descriptive Characteristics by RSM Trajectory Groups

|  |  | **RSM: Low-Decreasing** |  | **RSM: Mild-Decreasing** |  | **RSM: High Decreasing** |  |
| --- | --- | --- | --- | --- | --- | --- | --- |
|  |  | n=47 | m(SD) | n=98 | m(SD) | n = 48 | m(SD) |
| **Childhood IQ** | Verbal | 43 | 44.47 (20.89) | 87 | 34.74 (20.31) | 48 | 23.25 (13.57) |
|  | Nonverbal | 43 | 78.37 (19.75) | 87 | 65.78 (19.02) | 27 | 55.52 (20.86) |
| **Adulthood IQ** | Verbal | 26 | 88.35 (35.37) | 51 | 56.53 (44.29) | 26 | 27.08 (31.59) |
|  | Nonverbal | 37 | 92 (33.4) | 59 | 59.53 (39.78) | 14 | 31.12 (25.73) |
| **ADOS CSS Scores** | Childhood | 38 | 6.42 (2.62) | 86 | 7.49 (2.43) | 48 | 8.81 (1.32) |
|  | Adulthood | 24 | 4.83 (2.55) | 54 | 5.3 (2.12) | 17 | 4 (2.37) |
| **ADOS CSS Scores SA** | Childhood | 38 | 7.11 (2.53) | 86 | 7.84 (2.52) | 48 | 9 (1.35) |
|  | Adulthood | 25 | 5.6 (2.47) | 51 | 5.57 (2.29) | 20 | 4.55 (2.48) |
| **AMAS Score (age 18)** | Parent Report | 18 | 8.44 6.84 | 49 | 10.94 6.64 | 22 | 8.64 6.93 |
|  | Self Report | 11 | 15.82 8.9 | 14 | 13.64 8.23 | 2 | 10.5 0.71 |
| **AMAS Score (age 26)** | Parent Report | 22 | 9.09 7.06 | 51 | 11.57 6.06 | 22 | 7.36 6.15 |
|  | Self Report | 16 | 14.25 8.63 | 21 | 12.81 7.3 | N/A | N/A |
| **BDI Score (age 18)** | Parent Report | 23 | 9.14 9.96 | 53 | 8.08 7.79 | 19 | 10.47 7.6 |
|  | Self Report | 10 | 15.2 12.47 | 13 | 6.85 6.41 | N/A | N/A |
| **BDI Score (age 26)** | Parent Report | 20 | 4.35 9.64 | 49 | 5.51 6.66 | 23 | 4.13 5.16 |
|  | Self Report | 17 | 11.88 11.69 | 21 | 9.86 8.11 | N/A | N/A |
|  |  | n |  | n |  | n |  |
| **Anxiety Trajectory** | Stable-Low | 26 |  | 52 |  | 34 |  |
|  | Stable-High | 10 |  | 22 |  | 5 |  |
| **Depression Trajectory** | Stable-Low | 27 |  | 49 |  | 22 |  |
|  | High-Fluctuating | 9 |  | 26 |  | 17 |  |

**Table 3.** Descriptive Characteristics by IS Trajectory Groups

|  |  | Low-Increasing |  | High-Decreasing |  | *t* | df, n | *p* |
| --- | --- | --- | --- | --- | --- | --- | --- | --- |
| □ |  | n=162 | m (SD) | n=31 | m (SD) |  |  |  |
| Childhood IQ | Verbal | 147 | 33.56 (19.67) | 31 | 36 (23.17) | -0.6 | 1, 175 | 0.54 |
|  | Nonverbal | 147 | 66.65 (20.60) | 31 | 63.23 (24.20) | 0.81 | 1, 175 | 0.41 |
| Adulthood IQ | Verbal | 88 | 56.66 (44.16) | 15 | 59.87 (48.82) | -0.25 | 1, 100 | 0.79 |
|  | Nonverbal | 81 | 59.86 (41.30) | 15 | 62.33 (40.75) | -0.21 | 1, 93 | 0.83 |
| ADOS CSS Scores | Childhood | 143 | 7.64 (2.33) | 29 | 7.55 (2.60) | 0.17 | 1, 170 | 0.86 |
|  | Adulthood | 83 | 4.93 (2.32) | 12 | 5.08 (2.31) | -0.21 | 1, 93 | 0.82 |
| ADOS CSS Scores SA | Childhood | 143 | 8.07 (2.30) | 29 | 7.66 (2.59) | 0.86 | 1, 169 | 0.38 |
|  | Adulthood | 84 | 5.32 (2.41) | 12 | 5.67 (2.35) | -0.46 | 1, 93 | 0.64 |
| AMAS Score (age 18) | Parent Report | 77 | 9.74 (6.87) | 12 | 10.67 (6.41) | -0.43 | 1, 87 | 0.25 |
|  | Self Report | 23 | 14.87 (7.86) | 4 | 11 (10.36) | 0.87 | 1,25 | 0.39 |
| AMAS Score (age 26) | Parent Report | 81 | 9.8 (6.50) | 14 | 11.29 (6.59) | -0.78 | 1,92 | 0.43 |
|  | Self Report | 31 | 13.84 (7.66) | 7 | 11 (8.33) | 0.87 | 1,36 | 0.38 |
| BDI Score (age 18) | Parent Report | 82 | 8.57 (8.43) | 12 | 10.42 (7.18) | 0.517 | 1,92 | 0.47 |
|  | Self Report | 20 | 10.7 (10.52) | 4 | 6.75 (8.92) | 0.489 | 1,21 | 0.49 |
| BDI Score (age 26) | Parent Report | 79 | 4.9 (7.22) | 13 | 5 (6.12) | -0.047 | 1,89 | 0.96 |
|  | Self Report | 32 | 10.56 (10.14) | 7 | 10.14 (8.82) | 0.1 | 1, 36 | 0.91 |
|  |  | n = 124 |  | n = 26 |  | *X*<sup>2</sup> | df, n | *p* |
| Anxiety Trajectory | Stable-Low | 93 (75%) |  | 20 (77%) |  | 1.03E-29 | 1,147 | 1.00 |
|  | Stable-High | 31 (25%) |  | 6 (23%) |  |  |  |  |
| Depression Trajectory | Stable-Low | 83 (67%) |  | 15 (58%) |  | 0.45 | 1,147 | 0.50 |
|  | High-Fluctuating | 41 (33%) |  | 11 (42%) |  |  |  |  |

**Table 4.** Descriptive Characteristics by Verbal RRB Trajectory Groups

|  |  | **Low-Stable** |  | **High-Increasing** |  | *t* | **df, n** | *p* |
| --- | --- | --- | --- | --- | --- | --- | --- | --- |
|  |  | **n=68** | **m (SD)** | **n=59** | **m (SD)** |  |  |  |
| **Childhood IQ** | Verbal | 60 | 43.27 (21.62) | 51 | 38.8 (19.95) | 1.12 | 1, 109 | 0.26 |
|  | Nonverbal | 60 | 75.23 (19.48) | 51 | 71.24 (16.79) | 1.14 | 1, 109 | 0.25 |
| **Adulthood IQ** | Verbal | 40 | 85.75 (32.83) | 39 | 70.74 (39.53) | 1.83 | 1, 77 | 0.07 |
|  | Nonverbal | 33 | 90.15 (30.04) | 35 | 71.37 (34.88) | 2.37 | 1, 66 | 0.02 |
| **ADOS CSS Scores** | Childhood | 55 | 6.56 (2.64) | 51 | 7.29 (2.48) | -1.46 | 1, 104 | 0.14 |
|  | Adulthood | 38 | 5.11 (2.44) | 39 | 5.31 (2.08) | -0.39 | 1, 75 | 0.69 |
| **ADOS CSS Scores SA** | Childhood | 55 | 7 (2.63) | 51 | 7.57 (2.45) | -1.14 | 1, 104 | 0.25 |
|  | Adulthood | 37 | 5.57 (2.4) | 37 | 5.49 (2.21) | 0.15 | 1, 72 | 0.88 |

**Table 5.**
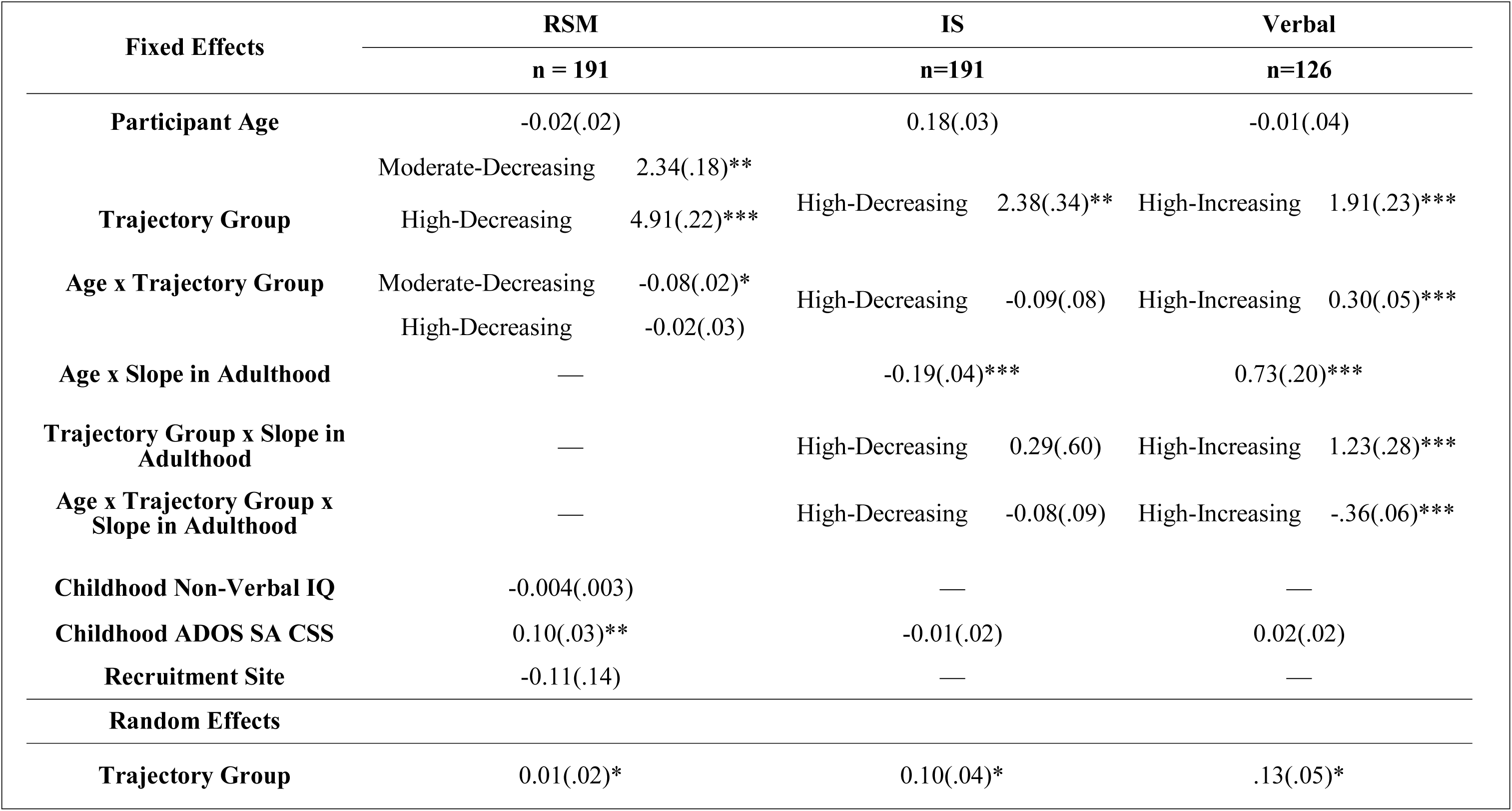
Rates of Growth in RSM, IS, and Verbal RRB Trajectories

| Fixed Effects | RSM |  | IS |  | Verbal |  |
| --- | --- | --- | --- | --- | --- | --- |
|  | n = 191 |  | n=191 |  | n=126 |  |
| **Participant Age** |  | -0.02(.02) |  | 0.18(.03) |  | -0.01(.04) |
| **Trajectory Group** | Moderate-Decreasing | 2.34(.18)** |  |  |  |  |
|  | High-Decreasing | 4.91(.22)*** | High-Decreasing | 2.38(.34)** | High-Increasing | 1.91(.23)*** |
| **Age x Trajectory Group** | Moderate-Decreasing | -0.08(.02)* |  |  |  |  |
|  | High-Decreasing | -0.02(.03) | High-Decreasing | -0.09(.08) | High-Increasing | 0.30(.05)*** |
| **Age x Slope in Adulthood** | — |  |  | -0.19(.04)*** |  | 0.73(.20)*** |
| **Trajectory Group x Slope in Adulthood** | — |  | High-Decreasing | 0.29(.60) | High-Increasing | 1.23(.28)*** |
| **Age x Trajectory Group x Slope in Adulthood** | — |  | High-Decreasing | -0.08(.09) | High-Increasing | -.36(.06)*** |
| **Childhood Non-Verbal IQ** |  | -0.004(.003) |  | — |  | — |
| **Childhood ADOS SA CSS** |  | 0.10(.03)** |  | -0.01(.02) |  | 0.02(.02) |
| **Recruitment Site** |  | -0.11(.14) |  | — |  | — |
| **Random Effects** |  |  |  |  |  |  |
| **Trajectory Group** |  | 0.01(.02)* |  | 0.10(.04)* |  | .13(.05)* |

Multilevel modeling via the mixed procedure in Stata was used to investigate differences in the intercepts and slopes (i.e., rates of change) of the various RSM, IS, and verbal trajectory subgroups. A multilevel modeling approach was selected for two reasons. First, because participants were nested across time points, multilevel modeling allowed us to account for the non-independence of assessment observations. Second, this approach enabled us to evaluate the influence of the inclusion of a random slope on trajectory estimates, and to allow for the possibility of differing slopes before and after age 9. Missing data were handled by estimating models using all available information. The models estimated included random slopes for trajectory group and fixed effects for age, trajectory group, slope after age 9, and age x trajectory group, age x slope, slope x trajectory group, and age x trajectory groups x slope interaction terms. All models were grand mean centered for age. Descriptive variables identified by one-way ANOVA and chi-square analyses as differing significantly between trajectory groups were included in the multilevel models as covariates. To account for multiple comparisons, Bonferroni corrections were used in interpreting all descriptive analyses.

## Results

### RRB Trajectory Analyses

#### RSM Trajectories

The best fitting model was a three-group model (Figure 1). The first trajectory group, Low-Decreasing, consisted of 28.2% of the sample. The second trajectory group, Moderate-Decreasing, consisting of 44.4% of the sample. Both the Low- and Moderate-Decreasing trajectories were best described by linear modeling. The third RSM trajectory group, High-Decreasing, was comprised of 27.3% of the sample and was best described by quadratic modeling. Compared to participants in the Low- and Moderate-Decreasing trajectory groups, those in the High-Decreasing RSM group had significantly lower childhood verbal IQ (F = 14.44; p <.001); childhood nonverbal IQ (F = 15.27; p <.001); adult verbal IQ (F = 15.82; p <.001); adult nonverbal IQ (F = 18.84; p <.001); and higher childhood ADOS overall CSS scores (F = 12.54; p = .001); as well as higher childhood ADOS SA CSS scores (F = 7.88; p = .001; Table 2).

##### Rate of Change in RSM Trajectories

Multilevel modeling was used to compare rates of change in the three RSM trajectory groups from ages 2-19. The inclusion of a main effect and interaction effects for slope did not significantly contribute to the RSM model and were thus dropped from the final model. There was also no significant effect of age in the RSM model (*p* = .294). However, there was a significant effect of group membership, with individuals in both the Moderate-Declining and High-Declining groups having significantly higher intercepts (i.e., RSM raw scores) than individuals in the Low-Declining group (both *p* < .001). There was also a significant age x trajectory group interaction, indicating inclusion in the Moderate-Declining group was associated with a significantly steeper decline in RSM raw scores between ages 2-19 than inclusion in the other two trajectory groups (*p* = .003). The inclusion of a trajectory group random-level slope significantly improved the fit of the model (*p* < .05), indicating that the rates of change in the three RRB Total trajectory groups significantly differed from one another. Finally, childhood ADOS SA CSS scores significantly contributed to the model (*p* = .001), though recruitment site (*p* = .61) and childhood nonverbal IQ (*p* = .24) did not.

#### IS Trajectories

A two-group model best fit the raw IS score trajectories. The first trajectory group, High-Decreasing, consisted of 81.5% of the sample, and the second group, Low-Increasing, consisted of 18.5% of the sample. The High-Decreasing group was best described by quadratic modeling, and the Low-Increasing group was best described by cubic modeling. There were no significant differences between descriptive characteristics of the two IS trajectory groups (Table 3).

##### Rate of Change in IS Trajectories

There was a significant effect of age in the IS model (*p* < .001) and a significant effect of group membership, indicating participants in the High-Increasing trajectory group had significantly higher IS raw scores than participants in the Low-Decreasing group (*p* < .001). The age x trajectory group, trajectory group x slope, and age x trajectory group x slope interaction terms were not significant. However, there was a significant slope x age interaction, indicating the rates of change for both the High-Increasing and the Low-Decreasing before age 9 were significantly different from the rates of change in both groups after age 9 (*p* < .001). In the Low-Decreasing group, IS scores were elevated at age 2 and increased slightly through age 9, then significantly declined from ages 9-19. In contrast, the High-Increasing group had low IS scores at age 2 and experienced a larger increase in IS scores from ages 2-9 than the Low-Decreasing group, followed by a plateau in IS scores from ages 9-19. The inclusion of random trajectory group level slopes also significantly improved the fit of the model (*p* < .05). Childhood ADOS SA CSS scores did not significantly contribute to the model (*p* = .59).

#### Verbal RRBs Trajectories

A two-group model was determined to be the best fit for the raw verbal score trajectories. The first trajectory group, High-Increasing, consisted of 50.7% of the sample, and the second group, Low-Stable, consisted of 49.3% of the sample. The High-Increasing group was best described by quadratic modeling, while the Low-Stable group was best described by intercept modeling. There were no significant differences in the descriptive characteristics of the two verbal RRB trajectory groups (Table 4).

##### Rate of Change in Verbal RRB Trajectories

There was no significant effect of age (*p* = .79) but a significant effect of group membership (*p* < .001) in the verbal model. There were also significant age x trajectory group (*p* < .001), and age x trajectory group x slope (*p* < .001) interaction terms. The inclusion of random trajectory group level slopes also significantly improved the fit of the model (*p* < .05). Childhood ADOS SA CSS scores did not significantly contribute to the model (*p* = .20). Verbal RRBs tended to increase with age, and participants in the High-Decreasing trajectory group had significantly higher verbal RRB raw scores than participants in the Low-Stable group. Rates of change in the verbal RRBs of the High-Decreasing and Low-Stable trajectory groups were significantly different from one another, with individuals in the High-Decreasing group experiencing a steeper rate of growth in verbal RRBs between ages 2-9. Verbal RRB raw scores increased in both trajectory groups over time, however, the scores of individuals in the High-Decreasing group increased at a faster rate from ages 2-9; the scores of both trajectory groups were stable from ages 9-19.

### RRB Trajectories and Internalizing Symptoms

The three RSM trajectory groups did not significant differ by likelihood of inclusion in the previously established depression (*p* = .23) or anxiety (*p* = .13) trajectory groups (McCauley et al., 2020). There were also no significant differences in parent-report AMAS scores (*p* = .25) and BDI scores (*p* = .54) nor self-report AMAS (*p* = .65) and BDI scores (*p* = .08) at age 19 between the three RSM Trajectory groups. Finally, there were no significant differences between the RSM trajectory groups in parent-report (*p* = .68) or self-report (*p* = .46) BDI scores or self-report AMAS scores (*p* = .73) at age 26 between the different RSM trajectory groups. However, parent-reported anxiety symptoms at age 26 were significantly higher amongst participants in the Moderate-Decreasing RSM trajectory group (F(2,92) = 3.70; *p* = .02) compared to participants in Low- and High-Decreasing groups (Table 2). This difference survived Bonferroni corrections.

The two IS trajectory groups did not significantly differ by likelihood of inclusion in the previously established depression (*p* = .36) or anxiety (*p* = .83) trajectory groups (McCauley et al., 2020). There were also no significant differences in parent-report AMAS scores (*p* = .25) and BDI scores (*p* = .470) nor self-report AMAS (*p* = .39) and BDI scores (*p* = .49) at age 19 between the two groups. There were also no significant differences between the IS trajectory groups in parent-report (*p* = .43) or self-report (*p* = .38) BDI scores nor parent-report (*p* = .96) or self-report AMAS scores (*p* = .91) at age 26 between the IS trajectory groups.

## Discussion

This study demonstrates that individuals with ASD experience different trajectories of RRB subtypes—specifically, restricted sensorimotor, insistence on sameness, and verbal restricted repetitive behaviors—from early childhood to young adulthood, as measured by the ADI-R. We identified three patterns of RSM trajectories, Low-Decreasing, Moderate-Decreasing, and High-Decreasing. IS trajectories followed High-Decreasing or Low-Increasing patterns. Finally, Verbal RRBs followed High-Increasing or Low-Stable trajectories. On average, symptoms of all three RRB subtypes significantly declined or plateaued by early adulthood. The current findings are in line with prior work in this sample examining trajectories of social communication symptoms (Bal et al., 2019) as well as work in separate samples examining developmental changes in core ASD symptoms (Gotham et al., 2012; Seltzer et al., 2004; Taylor & Seltzer, 2010) that found declines in core symptoms of ASD in early adulthood.

In contrast to some prior work (Baribeau et al., 2021a; Gotham et al., 2013, 2014; Lidstone et al., 2014; Rodgers et al., 2012; Stratis & Lecavalier, 2013) we found little evidence for associations between RRBs and internalizing symptoms in autistic adults. By longitudinally characterizing change and stability in RRB subtypes from ages 2-19, this paper adds to previous work in this sample (Richler et al., 2010) as well as work conducted in other samples ((Bishop et al., 2013; Courchesne et al., 2021; McDermott et al., 2020; Uljarević et al., 2021) that have found differing profiles of RRB symptoms in autistic individuals both within and across age groups. This study also corroborates findings from other longitudinal samples that, on average, core ASD symptomology decreases with increasing age (Seltzer et al., 2004; Taylor & Seltzer, 2010). Whereas prior work in this sample examined RRB trajectories using *a priori* groupings by diagnosis (ASD, PDD-NOS, or non-spectrum delays), the current study used a data-driven approach to categorize trajectory groupings.

For participants in the Low-Decreasing (28.2%) and Moderate-Decreasing (44.4%) RSM trajectory groups—almost three-quarters of the current sample—RSM behaviors were most pronounced in early childhood, between ages two and three, then declined through early adulthood (Figure 1). Individuals in the Moderate-Decreasing group had significantly higher RSM scores in early childhood than individuals in the Low-Decreasing group, however, both trajectories experienced similar negative linear growth across development, with the Moderate-Decreasing group’s RSM scores decreasing at a significantly faster rate than the Low-Decreasing group. In contrast, participants in the High-Decreasing group (27.3%) had more RSM symptoms in early childhood than participants in the other trajectory groups and experienced an increase in RSM symptoms across childhood, from ages 2-9. The High-Decreasing group’s RSM symptoms steadily declined in late childhood and adolescence such that by age 19, participants in the High-Decreasing group had similar levels of RSM symptoms as seen in early childhood.

Both IS trajectory groups, High-Decreasing (18.5%) and Low-Increasing (81.5%), experienced an increase in IS symptoms between ages 2-9, though the High-Decreasing group had significantly more IS symptoms at age 2 than the Low-Increasing group. The rate of growth for both trajectory groups significantly differed before and after age 9 (Figure 2), In contrast to the positive linear growth seen prior to age 9, IS behaviors decreased from 9-19 in the High-Decreasing group and plateaued during this same period in the Low-Increasing group. In part, insistence on sameness behaviors may increase from early to late childhood due to natural developmental changes associated with this life phase. For example, a two-year-old unhappy with a small change in the placement of a living room chair may not be able to effectively communicate their distress or physically move the chair back to its original “correct” placement. However, a 9-year-old could more easily convey their distress by complaining about the new position of the chair, attempting to move the chair back to the “correct” placement, or both—thus increasing the likelihood that their caregiver would endorse IS symptoms during the ADI-R. In this scenario, the child’s core IS symptomology may not necessarily change from early to late childhood, but their ability to make caregivers aware of these symptoms may increase markedly.

In addition to replicating and extending prior findings on RSM and IS symptom profiles in childhood and adulthood (Bishop et al., 2013; Evans et al., 2017; Guthrie et al., 2013; Richler et al., 2010), this study found evidence for distinct developmental trajectories of verbal RRBs. We identified two verbal RRB trajectory groups, Low-Stable (49.3%) and High-Increasing (50.7%). The rate of growth in verbal RRBs for the High-Increasing group significantly differed before and after age 9. The Low-Stable group displayed few to no verbal RRBs from ages 2-19; in contrast, the High-Increasing group experienced a sharp increase in verbal RRBs between ages 2-9, at which point their verbal RRBs plateaued. On average, participants in the High-Increasing group had higher verbal and non-verbal IQs than participants in the Low-Stable group. For autistic individuals with fluent speech, certain RRBs such as unusual preoccupations and circumscribed interests may be more apparent to caregivers, particularly if the autistic individual in question enjoys speaking about their restricted interests at length. In short, verbal RRBs appear to emerge in step with language fluency and stabilize as language fluency stabilizes.

A recent study of RRBs in minimally verbal children with ASD receiving intervention services found verbal RRBs such as echolalia, scripting and repetitive language and repetitive sounds increased during intervention (Harrop et al., 2021). Harrop et al. attributed this increase in verbal RRBs to increases in overall language production in response to intervention. Though the type of verbal RRBs an autistic individual presents with will vary based on their age, IQ, and verbal fluency, the current study complements and extends the findings of Harrop et al. (2021) by showing that RRBs such as circumscribed interests and unusual preoccupations are also directly tied to verbal ability.

The DSM-5 collapsed communication symptoms such as idiosyncratic phrases and echolalia into the RRB diagnostic criteria (APA, 2014). To reflect these DSM-5 inclusions to the categorization of RRBs, in addition to the RRB subtype totals summarized in Table 1, we also calculated RSM trajectory totals including raw item scores for stereotyped utterances and delayed echolalia and verbal RRB trajectory totals including raw item scores for stereotyped utterances, delayed echolalia and neologisms/idiosyncratic language. The inclusion of these additional items reflecting DSM-5 criteria did not change the fit of our RSM and Verbal RRB trajectory groups (Figure S2; Figure S3). The number of RSM and Verbal RRB trajectory groups and the patterns of growth of each trajectory group were not changed by the inclusion of scores from the stereotyped utterances, delayed echolalia and neologisms/idiosyncratic language ADI-R items. In other words, the inclusion of these additional symptoms in the DSM-5 criteria for RRB symptoms does not appear to alter the conceptual understanding of RRBs.

### RRBs and Internalizing Symptoms

We found little evidence of associations between RSM and IS behaviors and internalizing symptoms in adulthood. IS and RSM trajectory group membership was not associated with previously established trajectories in this sample of CBCL anxious/depressed T scores (McCauley et al., 2020), and neither parent- nor self-report anxiety and depression symptoms at age 19 or at age 26 significantly differed between the IS trajectory groups. Similarly, parent- and self-report depression and anxiety symptoms at age 19 were not related to RSM trajectory group membership. Parent-report—but not self-report—anxiety symptoms at age 26 were significantly higher for participants in the Moderate-Decreasing RSM trajectory group as compared to participants in the High-Decreasing RSM trajectory group. Though this difference survives Bonferroni corrections, it may still be reflective of Type II error and should be interpreted with caution.

These findings contrast with previous work that has found evidence for associations between IS and internalizing symptoms (Baribeau et al., 2021a; Gotham et al., 2013; Lidstone et al., 2014; Rodgers et al., 2012). The existing literature on associations between RRBs and internalizing symptomology has predominately focused on the experiences of verbal autistic individuals with average or better IQ (Gotham et al., 2013; Gotham et al., 2014). Associations between RRB symptomology and internalizing may be more pronounced in individuals with average or better IQ. Examinations of the relationship between IS and internalizing symptoms in a separate longitudinal sample of ASD, the Pathways in ASD cohort, have incorporated data from individuals with ASD of varying ability levels, but focused on associations between IS symptoms in early childhood and internalizing symptoms in early through late childhood (Baribeau et al., 2021). This contrasts with our study which looked at a longer period of development from early childhood through early adulthood. The current study also considered both parent- and self-report internalizing; Baribeau and colleagues used parent-report data only. It is also possible that the relationship between internalizing and IS changes across development. More specifically, it is possible that associations between insistence on sameness and anxiety may weaken with age especially for those who are less cognitively able and may differ by reporter. Furthermore, the Baribeau et al. study included additional items in their insistence on sameness factor such as Abnormal Response to Specific Sensory Stimuli; Sensitivity to Noise; and Circumscribed Interests. Their sample also had higher anxiety score trajectories than those in the current study, which may have also contributed to the differing findings.

### Limitations

This sample, followed longitudinally since initial evaluation in the early 1990’s, is comprised of a unique group of adults with ASD. These participants received an ASD diagnosis approximately 30 years ago, during an era when early diagnoses of autism were relatively uncommon. Thus, this sample may not be representative of people diagnosed with autism today. Attrition has reduced the number of African American participants and participants with low caregiver education in this cohort. This sample also includes relatively few female participants. Some prior work has found gender differences in RRBs (Frazier et al., 2014); the lack of females in our sample prevented us from examining potential gender differences in the current sample. Moreover, some measures of adult internalizing symptoms were parent-report. Though using parent-report measures allowed us to include participants who are minimally verbal, have high care needs, or were otherwise unable to independently complete measures of their internalizing symptoms in the current analyses, parent-report may not accurately reflect participants’ subjective symptomatology of anxiety and depression. However, for participants that were able to provide self-report BDI and AMAS data, neither self- nor parent-report depression and anxiety symptoms appeared to be strongly related to RRBs.

### Future Directions

Future studies should continue to examine patterns of RRB development into middle and later adulthood. Though research examining ASD symptoms and the experiences of autistic individuals and their families in early adulthood has increased in recent years, the field’s understanding of ASD in middle and later adulthood remains limited at best (Piven & Rabins, 2011). Prospectively monitoring how factors like gender and types of interventions and supports received affect long-term trajectories of core ASD symptoms also remains important.

Though this study found few associations between symptoms of depression and anxiety in adulthood and trajectories of RSM and IS behaviors, future work should consider examining relationships between these constructs, as well as how these relationships may vary across individuals with ASD of varying levels of ability across the life course. Internalizing symptoms may be more closely linked to RRBs, particularly IS, in autistic adults with average or better IQ (Gotham et al., 2013; Lidstone et al., 2014; Rodgers et al., 2012), but may not be related to RRBs in autistic adults with high needs. However, this is only one possible explanation of the current study’s null findings; further research is required to evaluate this. Future work should also attempt to identify methods of measuring internalizing symptoms in minimally verbal individuals with ASD that are not reliant on parent-report.

Understanding change and stability in core ASD symptomology throughout the life course is relevant to clinical practice. In treatment planning, understanding how a patient’s RRB symptomology may change over time can help clinicians and family members prioritize potential treatment targets. Further, developmental trajectories of RRBs may be relevant to applying ASD diagnostic criteria across the lifespan, as a young adult with autism may present differences in RRBs in symptom type, number, and intensity than a young child. The current study complements prior work in this sample characterizing stability in change in social communication symptoms of ASD from early childhood to adulthood (Bal et al., 2019).

### Conclusions

As increasing numbers of autistic individuals enter adulthood, understanding developmental changes in the presentation of ASD becomes increasingly important. The present study demonstrates variability in the trajectories of RSM, IS and Verbal restricted and repetitive behaviors from ages 2-19 as measured by the ADI-R, a caregiver-report clinician interview. These results provide insight into how RRBs may shift both within and across autistic individuals from early childhood into adulthood. Importantly, these findings illustrate that subtypes of RRBs show distinct developmental patterns. In the current sample, RSM behaviors declined from 2-19, with slightly less than one-third (27.3%) of participants experiencing an increase in RSM behaviors from 2-9, followed by a decrease from 9-19. In contrast, IS behaviors increased from ages 2-9, then declined or plateaued from 2-19, and Verbal RRBs increased from 2-9, then plateaued from 2-19—though 38.8% of the sample displayed few or no Verbal RRBs across development. ADOS Social Affect (SA) CSS scores from early childhood significantly contributed to RSM trajectories, with higher SA CSS scores associated with more RSM symptoms across development. However, SA CSS scores were not related to the IS and Verbal trajectory groups, and non-verbal IQ from early childhood was not related to change in the RSM, IS, or Verbal trajectories. This study failed to replicate prior findings relating IS to adult internalizing symptoms as measured by the CBCL, AMAS, and BDI but found preliminary evidence that a Mild-Decreasing pattern of RSM development may be related to anxiety in adulthood. Future work should continue to assess change and stability in RRB symptoms in autistic individuals through middle and later adulthood and examine relationships between RRB trajectories and other adult outcomes such as employment, living status, and subjective quality of life.

## Data Availability

All data produced in the present study are available upon reasonable request to the authors

## Acknowledgements

The authors would like to acknowledge the time participants and families have given to this project and thank them for their efforts. This study was funded by the National Institute of Child Health and Human Development R01 HD081199 (PI: CL), and the National Institute of Mental Health R01MH081873 (PI: CL)

## Notes

### Competing Interest Statement

Catherine Lord acknowledges the receipt of royalties from the sale of the Autism Diagnostic Observation Schedule-2 (ADOS-2) and the Autism Diagnostic Interview-Revised (ADI-R). Royalties generated from this study were donated to a not-for-profit agency, Have Dreams.

### Author Declarations

The IRB of UCLA gave ethical approval for this work.

## References

Achenbach, T. M., & Rescorla, L. A. (2001). Manual for the ASEBA School-Age Forms & Profiles. Burlington, VT: University of Vermont, Research Center for Children, Youth, & Families.

Achenbach, T. M., & Rescorla, L. A. (2003). Manual for the ASEBA Adult Forms & Profiles. Burlington, VT: University of Vermont, Research Center for Children, Youth, & Families.

American Psychiatric Association. (2014). Diagnostic and Statistical Manual of Mental Disorders: DSM-5 (5th edition) (Vol. 28). Emerald. https://doi.org/10.1108/rr-10-2013-0256

Anderson, D. K., Liang, J. W., & Lord, C. (2014). Predicting young adult outcome among more and less cognitively able individuals with autism spectrum disorders. Journal of Child Psychology and Psychiatry, and Allied Disciplines, 55(5), 485–494. https://doi.org/10.1111/jcpp.12178

Bal, V. H., Kim, S.-H., Fok, M., & Lord, C. (2019). Autism spectrum disorder symptoms from ages 2 to 19 years: Implications for diagnosing adolescents and young adults. Autism Research, 12(1), 89–99. https://doi.org/10.1002/aur.2004

Baribeau, D. A., Vigod, S., Pullenayegum, E., Kerns, C. M., Mirenda, P., Smith, I. M., Vaillancourt, T., Volden, J., Waddell, C., Zwaigenbaum, L., Bennett, T., Duku, E., Elsabbagh, M., Georgiades, S., Ungar, W. J., Zaidman-Zait, A., & Szatmari, P. (2020). Repetitive behavior severity as an early indicator of risk for elevated anxiety symptoms in autism spectrum disorder. Journal of the American Academy of Child & Adolescent Psychiatry, 59(7), 890–899.e3. https://doi.org/10.1016/j.jaac.2019.08.478

Baribeau, D. A., Vigod, S., Pullenayegum, E., Kerns, C. M., Mirenda, P., Smith, I. M., Vaillancourt, T., Volden, J., Waddell, C., Zwaigenbaum, L., Bennett, T., Duku, E., Elsabbagh, M., Georgiades, S., Ungar, W. J., Zait, A. Z., & Szatmari, P. (2021a). Co-occurring trajectories of anxiety and insistence on sameness behaviour in autism spectrum disorder. The British Journal of Psychiatry, 218(1), 20–27. https://doi.org/10.1192/bjp.2020.127

Baribeau, D. A., Vigod, S., Pullenayegum, E., Kerns, C. M., Mirenda, P., Smith, I. M., Vaillancourt, T., Volden, J., Waddell, C., Zwaigenbaum, L., Bennett, T., Duku, E., Elsabbagh, M., Georgiades, S., Ungar, W. J., Zait, A. Z., & Szatmari, P. (2021b). Co-occurring trajectories of anxiety and insistence on sameness behaviour in autism spectrum disorder. The British Journal of Psychiatry, 218(1), 20–27. https://doi.org/10.1192/bjp.2020.127

Beck, A. T., Steer, R. A., & Brown, G. K. (1996). Beck depression inventory-II. San Antonio, 78(2), 490–498.

Bishop, S. L., Hus, V., Duncan, A., Huerta, M., Gotham, K., Pickles, A., Kreiger, A., Buja, A., Lund, S., & Lord, C. (2013). Subcategories of restricted and repetitive behaviors in children with autism spectrum disorders. Journal of Autism and Developmental Disorders, 43(6), 1287–1297. https://doi.org/10.1007/s10803-012-1671-0

Boulter, C., Freeston, M., South, M., & Rodgers, J. (2014). Intolerance of uncertainty as a framework for understanding anxiety in children and adolescents with autism spectrum disorders. Journal of Autism and Developmental Disorders, 44(6), 1391–1402. https://doi.org/10.1007/s10803-013-2001-x

Carleton, R. N., Mulvogue, M. K., Thibodeau, M. A., McCabe, R. E., Antony, M. M., & Asmundson, G. J. G. (2012). Increasingly certain about uncertainty: Intolerance of uncertainty across anxiety and depression. Journal of Anxiety Disorders, 26(3), 468– 479. https://doi.org/10.1016/j.janxdis.2012.01.011

Courchesne, V., Bedford, R., Pickles, A., Duku, E., Kerns, C., Mirenda, P., Bennett, T., Georgiades, S., Smith, I. M., Ungar, W. J., Vaillancourt, T., Zaidman-Zait, A., Zwaigenbaum, L., Szatmari, P., Elsabbagh, M., & Pathways Team. (2021). Non-verbal IQ and change in restricted and repetitive behavior throughout childhood in autism: A longitudinal study using the Autism Diagnostic Interview-Revised. Molecular Autism, 12(1), 57. https://doi.org/10.1186/s13229-021-00461-7

Cuccaro, M. L., Shao, Y., Grubber, J., Slifer, M., Wolpert, C. M., Donnelly, S. L., Abramson, R. K., Ravan, S. A., Wright, H. H., DeLong, G. R., & Pericak-Vance, M. A. (2003). Factor analysis of restricted and repetitive behaviors in autism using the Autism Diagnostic Interview-R. Child Psychiatry and Human Development, 34(1), 3–17. https://doi.org/10.1023/a:1025321707947

Elliott, C. D. (2007). Differential Ability Scales (2nd ed.). Harcourt Assessment.

Esbensen, A. J., Seltzer, M. M., Lam, K. S. L., & Bodfish, J. W. (2009). Age-related differences in restricted repetitive behaviors in autism spectrum disorders. Journal of Autism and Developmental Disorders, 39(1), 57–66. https://doi.org/10.1007/s10803-008-0599-x

Evans, D. W., Uljarević, M., Lusk, L. G., Loth, E., & Frazier, T. (2017). Development of two dimensional measures of restricted and repetitive behavior in parents and children. Journal of the American Academy of Child & Adolescent Psychiatry, 56(1), 51–58. https://doi.org/10.1016/j.jaac.2016.10.014

Frazier, T. W., Georgiades, S., Bishop, S. L., & Hardan, A. Y. (2014). Behavioral and cognitive characteristics of females and males with autism in the Simons Simplex Collection. Journal of the American Academy of Child and Adolescent Psychiatry, 53(3), 329–340.e3. https://doi.org/10.1016/j.jaac.2013.12.004

Georgiades, S., Papageorgiou, V., & Anagnostou, E. (2010). Brief report: Repetitive behaviours in Greek individuals with autism spectrum disorder. Journal of Autism and Developmental Disorders, 40(7), 903–906. https://doi.org/10.1007/s10803-009-0927-9

Gotham, K., Bishop, S. L., Brunwasser, S., & Lord, C. (2014). Rumination and perceived impairment associated with depressive symptoms in a verbal adolescent-adult ASD sample. Autism Research, 7(3), 381–391. https://doi.org/10.1002/aur.1377

Gotham, K., Bishop, S. L., Hus, V., Huerta, M., Lund, S., Buja, A., Krieger, A., & Lord, C. (2013). Exploring the relationship between anxiety and insistence on sameness in autism spectrum disorders. Autism Research, 6(1), 33–41. https://doi.org/10.1002/aur.1263

Gotham, K., Pickles, A., & Lord, C. (2012). Trajectories of autism severity in children using standardized ADOS scores. Pediatrics, 130(5), e1278–e1284.

Guthrie, W., Swineford, L. B., Nottke, C., & Wetherby, A. M. (2013). Early diagnosis of autism spectrum disorder: Stability and change in clinical diagnosis and symptom presentation. Journal of Child Psychology and Psychiatry, 54(5), 582–590. https://doi.org/10.1111/jcpp.12008

Harrop, C., Sterrett, K., Shih, W., Landa, R., Kaiser, A., & Kasari, C. (2021). Short-term trajectories of restricted and repetitive behaviors in minimally verbal children with autism spectrum disorder. Autism Research, 14(8), 1789–1799. https://doi.org/10.1002/aur.2528

Hedley, D., Uljarević, M., Wilmot, M., Richdale, A., & Dissanayake, C. (2018). Understanding depression and thoughts of self-harm in autism: A potential mechanism involving loneliness. Research in Autism Spectrum Disorders, 46, 1–7. https://doi.org/10.1016/j.rasd.2017.11.003

Hiruma, L., Pretzel, R. E., Tapia, A. L., Bodfish, J. W., Bradley, C., Wiggins, L., Hsu, M., Lee, L.-C., Levy, S. E., & Daniels, J. (2021). A distinct three-factor structure of restricted and repetitive behaviors in an epidemiologically sound sample of preschool-age children with autism spectrum disorder. Journal of Autism and Developmental Disorders. https://doi.org/10.1007/s10803-020-04776-x

Hollocks, M. J., Lerh, J. W., Magiati, I., Meiser-Stedman, R., & Brugha, T. S. (2018). Anxiety and depression in adults with autism spectrum disorder: A systematic review and meta-analysis. Psychological Medicine, 49(4), 559–572. https://doi.org/10.1017/s0033291718002283

Hu, H.-F., Liu, T.-L., Hsiao, R. C., Ni, H.-C., Liang, S. H.-Y., Lin, C.-F., Chan, H.-L., Hsieh, Y.-H., Wang, L.-J., Lee, M.-J., Chou, W.-J., & Yen, C.-F. (2019). Cyberbullying victimization and perpetration in adolescents with high-functioning autism spectrum disorder: correlations with depression, anxiety, and suicidality. Journal of Autism and Developmental Disorders, 49(10), 4170–4180. https://doi.org/10.1007/s10803-019-04060-7

Jones, R. M., Pickles, A., & Lord, C. (2017). Evaluating the quality of peer interactions in children and adolescents with autism with the Penn Interactive Peer Play Scale (PIPPS). Molecular Autism, 8, 28. https://doi.org/10.1186/s13229-017-0144-x

Lai, M.-C., Kassee, C., Besney, R., Bonato, S., Hull, L., Mandy, W., Szatmari, P., & Ameis, S. H. (2019). Prevalence of co-occurring mental health diagnoses in the autism population: A systematic review and meta-analysis. The Lancet Psychiatry, 6(10), 819– 829. https://doi.org/10.1016/S2215-0366(19)30289-5

Lam, K. (2004). The Repetitive Behavior Scale-Revised: Independent validation and the effects of subject variables. The Ohio State University.

Lam, K., Bodfish, J. W., & Piven, J. (2008). Evidence for three subtypes of repetitive behavior in autism that differ in familiality and association with other symptoms. Journal of Child Psychology and Psychiatry, and Allied Disciplines, 49(11), 1193–1200. https://doi.org/10.1111/j.1469-7610.2008.01944.x

Le Couteur, A., Rutter, M., Lord, C., Rios, P., Robertson, S., Holdgrafer, M., & McLennan, J. (1989). Autism diagnostic interview: A standardized investigator-based instrument. Journal of Autism and Developmental Disorders, 19(3), 363–387. https://doi.org/10.1007/BF02212936

Lidstone, J., Uljarević, M., Sullivan, J., Rodgers, J., McConachie, H., Freeston, M., Le Couteur, A., Prior, M., & Leekam, S. (2014). Relations among restricted and repetitive behaviors, anxiety and sensory features in children with autism spectrum disorders. Research in Autism Spectrum Disorders, 8(2), 82–92. https://doi.org/10.1016/j.rasd.2013.10.001

Lord, C., Risi, S., DiLavore, P. S., Shulman, C., Thurm, A., & Pickles, A. (2006). Autism from 2 to 9 years of age. Archives of General Psychiatry, 63(6), 694–694. https://doi.org/10.1001/archpsyc.63.6.694

Lord, C., Rutter, M., DiLavore, P. C., Risi, S., Gotham, K., & Bishop, S. (2012). Autism diagnostic observation schedule: ADOS-2. Western Psychological Services Torrance.

Lowe, P. A., & Reynolds, C. R. (2004). Psychometric analyses of the adult manifest anxiety scale–adult version among young and middle-aged adults. Educational and Psychological Measurement, 64(4), 661–681. https://doi.org/10.1177/0013164404263881

Lugo-Marín, J., Magán-Maganto, M., Rivero-Santana, A., Cuellar-Pompa, L., Alviani, M., Jenaro-Rio, C., Díez, E., & Canal-Bedia, R. (2019). Prevalence of psychiatric disorders in adults with autism spectrum disorder: A systematic review and meta-analysis. Research in Autism Spectrum Disorders, 59, 22–33. https://doi.org/10.1016/j.rasd.2018.12.004

Magiati, I., Tay, X. W., & Howlin, P. (2014). Cognitive, language, social and behavioural outcomes in adults with autism spectrum disorders: A systematic review of longitudinal follow-up studies in adulthood. Clinical Psychology Review, 34(1), 73–86. https://doi.org/10.1016/J.CPR.2013.11.002

McCauley, J. B., Elias, R., & Lord, C. (2020). Trajectories of co-occurring psychopathology symptoms in autism from late childhood to adulthood. Development and Psychopathology, 32(4), 1287–1302. https://doi.org/10.1017/S0954579420000826

McDermott, C. R., Farmer, C., Gotham, K. O., & Bal, V. H. (2020). Measurement of subcategories of repetitive behaviors in autistic adolescents and adults. Autism in Adulthood, 2(1), 48–60.

Mirenda, P., Smith, I. M., Vaillancourt, T., Georgiades, S., Duku, E., Szatmari, P., Bryson, S., Fombonne, E., Roberts, W., & Volden, J. (2010). Validating the repetitive behavior scale-revised in young children with autism spectrum disorder. Journal of Autism and Developmental Disorders, 40(12), 1521–1530.

Moore, V., & Goodson, S. (2003). How well does early diagnosis of autism stand the test of time? Follow-up study of children assessed for autism at age 2 and development of an early diagnostic service. Autism: The International Journal of Research and Practice, 7(1), 47–63. https://doi.org/10.1177/1362361303007001005

Mullen, E. M. (1995). Mullen Scales of Early Learning (AGS Edition). American Guidance Service. https://doi.org/10.1007%2F978-0-387-79948-3_1570

Nagin, D. S., Jones, B. L., Passos, V. L., & Tremblay, R. E. (2018). Group-based multi-trajectory modeling. Statistical Methods in Medical Research, 27(7), 2015–2023. https://doi.org/10.1177/0962280216673085

Pickles, A., McCauley, J. B., Pepa, L. A., Huerta, M., & Lord, C. (2020). The adult outcome of children referred for autism: Typology and prediction from childhood. Journal of Child Psychology and Psychiatry, n/a(n/a). https://doi.org/10.1111/jcpp.13180

Piven, J., & Rabins, P. (2011). Autism spectrum disorders in older adults: toward defining a research agenda. Journal of the American Geriatrics Society, 59(11), 2151–2155. https://doi.org/10.1111/j.1532-5415.2011.03632.x

Reynolds, C. R., Richmond, B. O., & Lowe, P. A. (2003). The Adult Manifest Anxiety Scale (AMAS): Manual. Western Psychological Services (WPS).

Richler, J., Bishop, S. L., Kleinke, J. R., & Lord, C. (2007). Restricted and repetitive behaviors in young children with autism spectrum disorders. Journal of Autism and Developmental Disorders, 37(1), 73–85. https://doi.org/10.1007/s10803-006-0332-6

Richler, J., Huerta, M., Bishop, S. L., & Lord, C. (2010). Developmental trajectories of restricted and repetitive behaviors and interests in children with autism spectrum disorders. Development and Psychopathology, 22(1), 55–69. https://doi.org/10.1017/S0954579409990265

Rodgers, J., Glod, M., Connolly, B., & McConachie, H. (2012). The relationship between anxiety and repetitive behaviours in autism spectrum disorder. Journal of Autism and Developmental Disorders, 42(11), 2404–2409. https://doi.org/10.1007/s10803-012-1531-y

Rutter, M., LeCouteur, A., & Lord, C. (2003). Autism Diagnostic Interview-Revised (ADI-R). Western Psychological Services.

Seltzer, M. M., Shattuck, P., Abbeduto, L., & Greenberg, J. S. (2004). Trajectory of development in adolescents and adults with autism. Mental Retardation and Developmental Disabilities Research Reviews, 10(4), 234–247.

Stratis, E. A., & Lecavalier, L. (2013). Restricted and repetitive behaviors and psychiatric symptoms in youth with autism spectrum disorders. Research in Autism Spectrum Disorders, 7(6), 757–766. https://doi.org/10.1016/j.rasd.2013.02.017

Taylor, J. L., & Seltzer, M. M. (2010). Changes in the autism behavioral phenotype during the transition to adulthood. Journal of Autism and Developmental Disorders, 40(12), 1431–1446.

Uljarević, M., Frazier, T. W., Jo, B., Billingham, W. D., Cooper, M. N., Youngstrom, E. A., Scahill, L., & Hardan, A. Y. (2021). Big data approach to characterize restricted and repetitive behaviors in autism. Journal of the American Academy of Child and Adolescent Psychiatry, S0890-8567(21)01317-4. https://doi.org/10.1016/j.jaac.2021.08.006

van Steensel, F. J. A., & Heeman, E. J. (2017). Anxiety levels in children with autism spectrum disorder: a meta-analysis. Journal of Child and Family Studies, 26(7), 1753–1767. https://doi.org/10.1007/s10826-017-0687-7

Volden, J., & Lord, C. (1991). Neologisms and idiosyncratic language in autistic speakers. Journal of Autism and Developmental Disorders, 21(2), 109–130. https://doi.org/10.1007/BF02284755

Wechsler, D. (1999). The Wechsler abbreviated scale for intelligence. The Psychological Corporation.

Wurzman, R., Forcelli, P. A., Griffey, C. J., & Kromer, L. F. (2015). Repetitive grooming and sensorimotor abnormalities in an ephrin-a knockout model for autism spectrum disorders. Behavioural Brain Research, 278, 115–128. https://doi.org/10.1016/j.bbr.2014.09.012

