## Supplemental Tables and Figures for "Development of Restricted and Repetitive Behaviors from 2-19 and Internalizing Symptom Outcomes in a Longitudinal Study of Autism"

**Supplementary Table/Figure Legends**

Supplementary Tables

*Supplementary Table 1.* Descriptive Characteristics of Internalizing Symptoms Subsample

Supplementary Figures

*Supplemental Figure 1.* Trajectories Of Raw Verbal RRB Scores Including Verbal And Non-verbal/minimally Participants.

*Supplemental Figure 2.* Trajectories Of Raw Verbal RRB Scores Including Additional Items: Stereotyped Utterances and Delayed Echolalia; and Neologisms/Idiosyncratic Language

*Supplemental Figure 3.* Trajectories Of Raw RSM Scores Including Additional Item: Stereotyped Utterances and Delayed Echolalia

**Table S1.** Descriptive Characteristics of Internalizing Symptoms Subsample

|  |  | **Current Subsample (*n* = 154)** | **Total Sample (N = 39)** |  |
| --- | --- | --- | --- | --- |
| **Participant Race** |  | 114 | 21 | *X*^2^(1, 193) = 6.64, *p =.084* |
|  | White |  |  |  |
|  | Black | 37 | 17 |  |
|  | Asian | 2 | 1 |  |
|  | American Indian | 0 | 0 |  |
|  | Multiracial | 1 | 0 |  |
| **Participant Gender** | Male | 134 | 33 | *X^2^(1, 193) = .153, p =.69* |
|  | Female | 20 | 6 |  |
| **Caregiver Education** | < Four-year degree | 126 | 25 | *X^2^(2, 193) = 5.73 p =0.016* |
|  | > Four-year degree | 28 | 14 |  |
| **Site** | North Carolina | 77 | 26 | X^2^(2, 196) = 3.48, p =.1749 |
|  | Chicago | 64 | 11 |  |
|  | Michigan | 13 | 2 |  |
| **First Non-Verbal IQ**^a^ | M(SD) | 68.84 (22.46) | 61.41 (21.36) | *t*(191) = -1.86, *p* = .063 |
| **First Verbal IQ**^a^ | M(SD) | 39.85 (26.61) | 31.35 (19.95) | *t*(191) = -1.86, *p* = 0.0  6 |

**Figure S1.** Trajectories Of Raw Verbal RRB Scores Including Verbal And Non-verbal/minimally Participants.

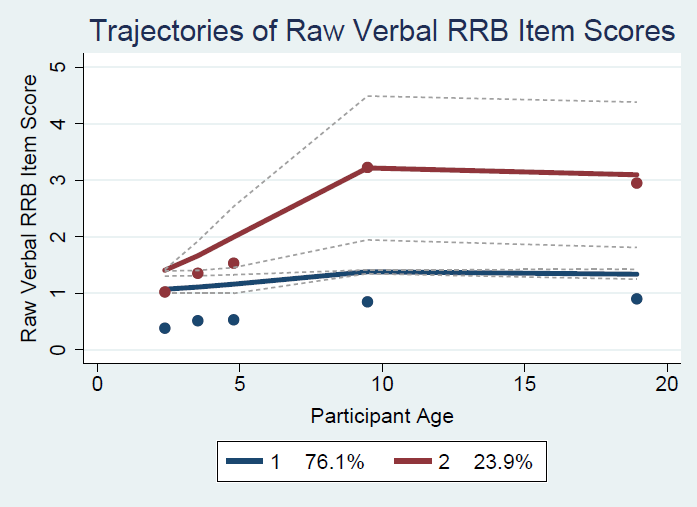

**Figure S2.** Trajectories Of Raw Verbal RRB Scores Including Additional Items: Stereotyped Utterances and Delayed Echolalia; and Neologisms/Idiosyncratic Language

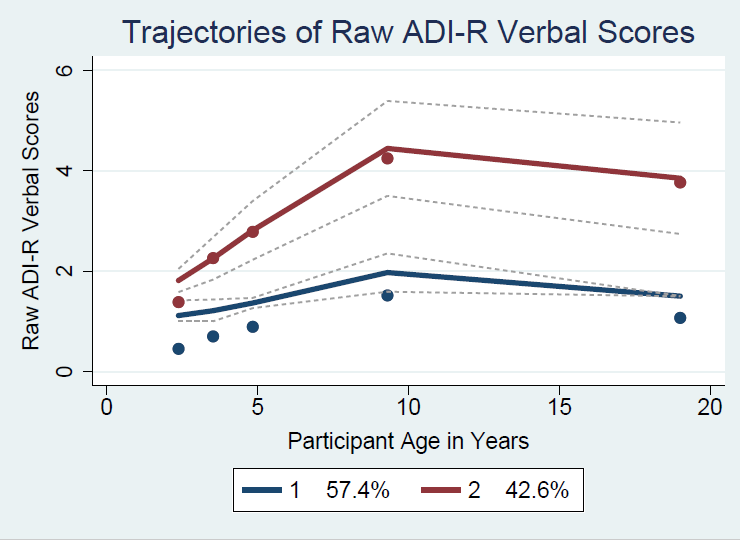

**Figure S3.** Trajectories Of Raw RSM Scores Including Additional Item: Stereotyped Utterances and Delayed Echolalia

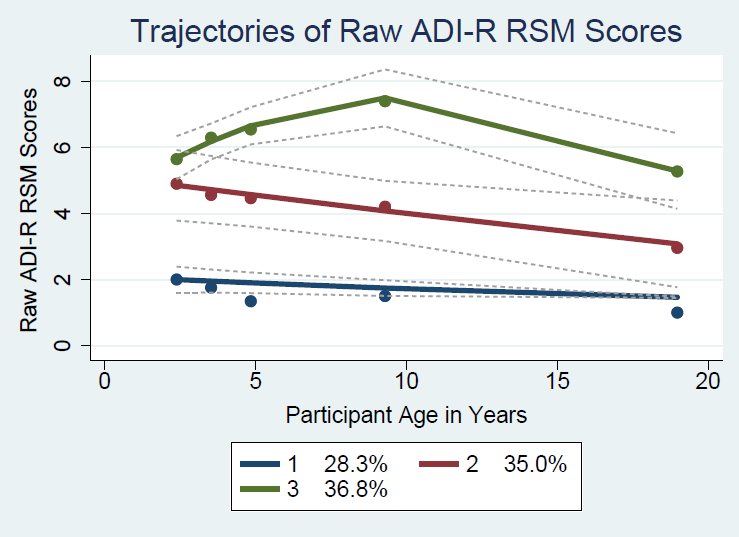
